# Estimation of the SARS-CoV-2 infection fatality rate in Germany

**DOI:** 10.1101/2021.01.26.21250507

**Authors:** Thomas Dimpfl, Jantje Sönksen, Ingo Bechmann, Joachim Grammig

**Author notes:** Corresponding authors: Thomas Dimpfl, Jantje Sönksen, Joachim Grammig, University of Tübingen, Department of Statistics and Econometrics, Sigwartstr. 18, 72076 Tübingen, Germany. These authors share first co-authorship.

## Abstract

Assessing the infection fatality rate (IFR) of SARS-CoV-2 is one of the most controversial issues during the pandemic. Due to asymptomatic or mild courses of COVID-19, many infections remain undetected. Reported case fatality rates – COVID-19-associated deaths divided by number of detected infections – are therefore poor estimates of the IFR. Endogenous changes of the population at risk of a SARS-CoV-2 infection, changing test practices and an improved understanding of the pathogenesis of COVID-19 further exacerbate the estimation of the IFR. Here, we propose a strategy to estimate the IFR of SARS-CoV-2 in Germany that combines official data on reported cases and fatalities supplied by the Robert Koch Institute (RKI) with data from seroepidemiological studies in two infection hotspots, the Austrian town Ischgl and the German municipality Gangelt, respectively. For this purpose, we use the law of total probability to derive an approximate formula for the IFR that is based on a set of assumptions regarding data quality and test specificity and sensitivity. The resulting estimate of the IFR in Germany of 0.83% (95% CI: [0.69%; 0.98%]) that is based on a combination of the RKI and Ischgl data is notably higher than the IFR estimate reported in the Gangelt study (0.36% [0.29%; 0.45%]). It is closer to the consolidated estimate based on a meta-analysis (0.68% [0.53%; 0.82%]), where the difference can be explained by Germany’s disadvantageous age structure. As a result of virus mutations, vaccination strategies, and improved therapy, a re-estimation of the IFR will eventually be mandated; the proposed method is able to account for such developments.

## Introduction

Estimating the infection fatality rate (IFR) of SARS-CoV-2 as it applies to the population of a country (here: Germany) by using official data on detected infections and case fatalities (here: data provided by the Robert Koch Institute, RKI) is hampered for two main reasons. First, the information contained in the reported SARS-CoV-2 infection data depends on the country’s testing strategy, which changes in the course of the pandemic.^1, 2^ Because capacities are limited, testing must be, in particular at the beginning of the pandemic, prioritized for symptomatic persons and their contacts. As a result, many SARS-CoV-2 infections remain undetected and unreported.^3–5^ Younger individuals without pre-existing conditions who become infected are often asymptomatic or show mild courses of COVID-19.^6, 7^ For this subpopulation, a much smaller or even negligible IFR applies, and they will be initially underrepresented in the subpopulation of those who tested positive for a SARS-CoV-2 infection. As capacities increase, an extended testing strategy will also capture asymptomatic infected persons, such that the composition of the subpopulation of infected and positively tested changes, including also younger individuals without pre-existing conditions.^8^ Second, as the pandemic evolves, members of high-risk subpopulations (elderly and individuals with pre-existing conditions) will try to protect themselves from a SARS-CoV-2 infection, either by voluntary self-isolation and social distancing or receiving vaccination. They will also benefit from lockdown measures, and may be offered special protection (e.g., in nursery homes).^9^ Accordingly, the country’s population at risk of a SARS-CoV-2 infection changes endogenously. As members of the high-risk subpopulations effectively avoid a SARS-CoV-2 infection, the composition of the infected subpopulation and that of the reported cases change, too.

Both effects, the endogenous change of the composition of the population at risk of a SARS-CoV-2 infection and the modifications of the testing strategy, tend to decrease the reported case fatality rates. This development is depicted in Fig. 1, which shows, using RKI data, the daily updated estimted CFRs in Fig. 1a (details of the computation are provided below), along with reported new cases (Fig. 1b), COVID-19-associated deaths (Fig. 1c), and number of tests (Fig. 1d).

**Fig. 1.**
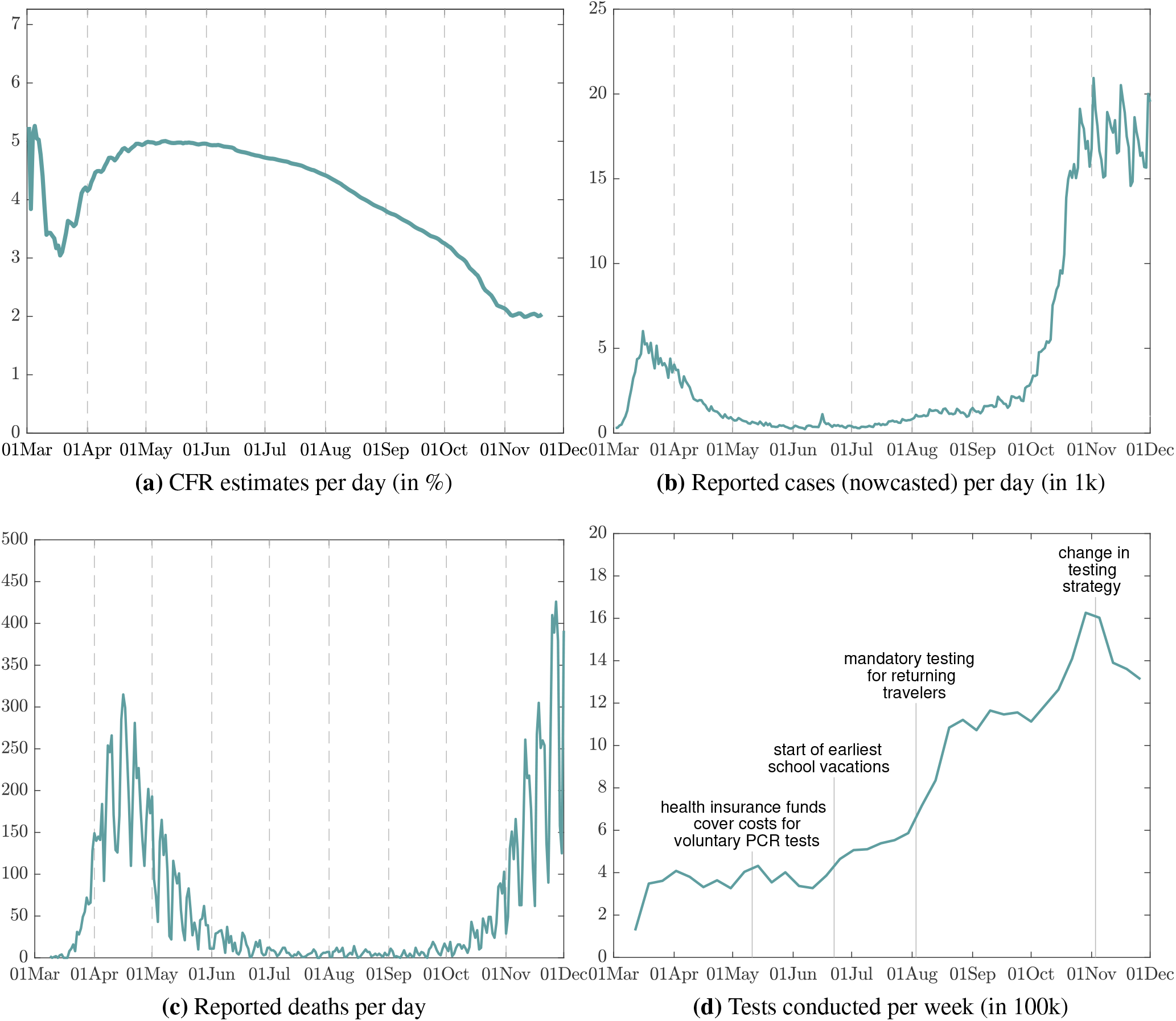
CFR estimates, nowcast of reported infections, COVID-19-associated deaths, and SARS-CoV-2 PCR tests in Germany. The figure shows in Panel 1a the CFR estimates, which are obtained as the ratio of COVID-19-associated fatalities shifted backwards by 19 days and the RKI nowcast of the number of reported infections per day. The latter are depicted in Panel 1b. Panel 1c depicts the number of COVID-19-associated deaths per day (not backward-shifted). Panel 1d shows the number of tests conducted per week. All raw data are provided by the RKI.

Seroepidemiological studies of infection hotspots are less prone to the described sample selection effects. Ideally, the entire hotspot population or a random sample is tested by means of a serological test for SARS-CoV-2 antibodies as well as a study-site PCR test. A drawback, from a statistical perspective, is that the number of COVID-19-associated fatalities recorded in the hotspot studies will be small. For example, in the seroepidemiological study conducted for the Austrian hotspot Ischgl,^10^ only two deaths associated with COVID-19 are reported. However, the two seroepidemiological studies that we use also provide information about whether a SARS-CoV-2 infection had been detected by prior (selective) PCR testing or not. This information is an important input for the IFR estimation strategy described in the following.

### Estimation strategy and data

The proposed strategy to estimate the IFR of SARS-CoV-2 in Germany combines the country-wide RKI data on COVID-19-associated fatalities and reported SARS-CoV-2 cases with the data obtained from the seroepidemiological studies for the two infection hotspots Ischgl (Austria)^10^ and Gangelt (Germany).^11^ The approach exploits a link of IFR and CFR that can be established by involving the law of total probability, which is detailed in the Methods section. It is based on a set of assumptions regarding the statistical properties of the SARS-CoV-2 PCR test used to assess the country-wide number of infections and the serological tests in the hotspot study. In particular, it is assumed that the selective SARS-CoV-2 PCR test has positive predictive power (PPV) and sensitivity close to 100%. Moreover, it is assumed that the study-site serological test has a specificity close to 100%. In addition, it is assumed that all COVID-19-associated deaths are detected and reported. In Germany, severe cases of a SARS-CoV-2 infection that require hospitalization will not go undetected, so that it is reasonable to assume that the RKI data account for all fatalities associated with COVID-19.

The IFR is defined here as P(M = 1| C = 1), the probability that a person infected with SARS-CoV-2 (C = 1, else C = 0) dies of/with COVID-19 (M = 1, else M = 0). If the aforementioned assumptions hold, then, as formally shown in the Methods section, the IFR can be approximated as follows:

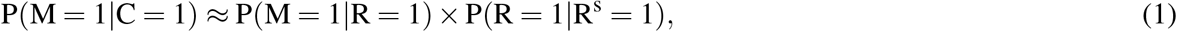

where P(M = 1 | R = 1) is the case fatality rate (CFR). It denotes the probability that a person who tested PCR-positive (R = 1) dies of/with COVID-19. In addition, P(R = 1 | R^s^ = 1) denotes the probability that a person who tests positive in a seroepidemiological study (R^s^ = 1, else R^s^ = 0) has previously tested PCR-positive in the course of the selective testing strategy. It is referred to the detection probability, because it is approximately identical to P(R = 1| C = 1) if the aforementioned assumptions hold. The advantage of the linking formula (1) is that the two components on the right-hand side, the CFR and the detection probability, can be estimated using available data, which circumvents the problem of missing observations on SARS-CoV-2-infected persons. An estimate of the IFR is thus obtained by multiplying the CFR estimate with the estimated detection probability.

The estimation of the CFR is based on the RKI data of newly reported SARS-CoV-2 cases per day. The data refer to the subpopulation of individuals that have tested positive by means of a PCR test. Assuming that the selective PCR test has a positive predictive value of 100% this subpopulation is identical with the infected and positively tested subpopulation.^12, 13^ The RKI applies a nowcast to associate each case with the most likely infection date. The nowcast data are provided starting March 2, 2020. The RKI also supplies data on newly reported COVID-19-associated fatalities at the daily frequency. To align reported cases and COVID-19-associated deaths, we account for an incubation period of 5 days^14, 15^ and allow for an average of 14 days between the first time when symptoms occur until death.^16, 17^ Accordingly, we use a 19 days backward shift to align the fatality data with the RKI’s nowcast of infections.^18^ Dividing the backward-shifted fatalities by the number of nowcasted infections over a selected period of time provides an estimate of the CFR. The CFR estimate thus obtained can be updated at a daily frequency as shown in Fig. 1a.

To estimate the detection probability P(R = 1 | R^s^ = 1), the second component of the IFR formula in Eq. (1), we exploit information contained in the seroepidemiological studies conducted in Ischgl^10^ (April 21 - 27, 2020) and in Gangelt (March 31 - April 6, 2020).^11^ The Gangelt study uses a random sample of the municipality’s population, while in Ischgl all inhabitants were invited to join the study with a participation rate of 79%. The estimation becomes possible, because both studies report in detail whether an individual is seropositive, tested positive in a study-site PCR test, and whether the person had a prior positive PCR test result. Drawing on the these data for the present analysis, the Ischgl study offers three advantages. The first is a larger sample size. The number of individuals with an evaluable infection status is 1,473 in Ischgl compared to 919 in Gangelt. Second, due to a combination of tests, the authors of the Ischgl study achieve a specificity of the combined serological testing procedure of 100%, which is favorable for the IFR approximation formula (1). Third, the number of individuals who tested study-site PCR-positive is small. show below, the decision how to resolve the uncertainty whether to count the study-site PCR test positives as cases that would have remained undetected without the study-site test or not does not matter much. For study participants with a positive study-site PCR test, we cannot know whether their infection status would have gone undetected had they not taken part in the study. The Gangelt study reports, in absolute and relative terms, a larger number of study-site SARS-CoV-2-positive PCR tests. Here, the decision how to resolve the uncertainty regarding detection matters more. Possible concerns to link German (RKI) and Austrian data are mitigated, because of the similar health systems of the two countries^19^, their demographics, and the fact that neither health system was overwhelmed during the pandemic.

When combining the RKI data and the data from the seroepidemiological studies in the proposed way, the following conditions must be met: First, the country-wide and hotspot testing strategies (initial, not study-site) must be comparable, because they determine the composition of the subpopulation of infected and positively tested individuals, which are assumed to match. Second, the composition of the populations at risk of a SARS-CoV-2 infection must be comparable, country-wide and in the infection hotspot. It is also assumed that the country-wide sample and the hotspot sample are independently drawn, an unproblematic assumption when combining the RKI and Ischgl data. These caveats mandate that the country-wide data used to estimate the CFR and the hotspot data on which the estimation of the detection probability is based should come from a time period during which these compatibility conditions can be defended.

## Results

To compute the IFR estimate according to the described strategy, it is necessary to first determine the time period for which the RKI data used to estimate the CFR are extracted. The base case uses data on RKI nowcasted cases and backward-shifted fatalities from March 2 until May 1, 2020. A robustness check varies the end date from April 1 to June 1. This time period is selected for the following considerations: First, it implies that the estimation of the CFR can be based on a sufficient number of cases (161,880 PCR-positive individuals by May 1, 2020) and fatalities (8,059 COVID-19-associated deaths by May 1, 2020). Second, the selected time range corresponds to the period within which the two seroepidemiological studies in Gangelt and Ischgl were conducted. Third, no notable changes of the country-wide testing strategy occurred during the selected time period (see Fig. 1d). Fourth, the assumption that the composition of the population at risk of a SARS-CoV-2 infection remains stable during the selected time interval seems reasonable. Finally, the choice mitigates seasonal effects of infections and disease.

Fig. 1a depicts the estimated CFRs, which for illustration purposes also set later end dates of the time interval used to count cases and fatalities. Note that at the onset of the pandemic, the CFR calculation must be based on a very small number of cases and fatalities. As SARS-CoV-2 infections and COVID-19-associated deaths grow during the first wave, the CFR estimates stabilize by the beginning of April. The decrease that sets in after June 1, 2020 reflects the aforementioned effects of increased test capacities and changing composition of the population at risk.

The base case estimate of the detection probability, the second component of the IFR formula above, draws on information contained in the seroepidemiological study conducted in Ischgl. As mentioned supra, one of the advantages of the Ischgl study is that there are fewer current infections detected by the study-site PCR test. In particular, the authors report that this test revealed eight new positive cases, five of which were also seropositive, while three were not (see also Table 1).^10^ Regarding these new PCR-positive cases, an assumption has to be made whether the SARS-CoV-2 infection would have been detected without the study-site PCR test or not. If the individuals were asymptomatic, the infection likely would have gone unnoticed. An alternative strategy is to exclude all study-site PCR-positive cases cases from the analysis, but we prefer another one, which provides the base case for the present analysis. This strategy excludes all study-site PCR-positive individuals who are not (yet) seropositive. However, it retains the study-site PCR-positive and seropositive cases, assuming that their infection would not have been detected without the study-site PCR test. Table 1 shows five more variants and the associated estimates of the detection probability.

**Table 1.**
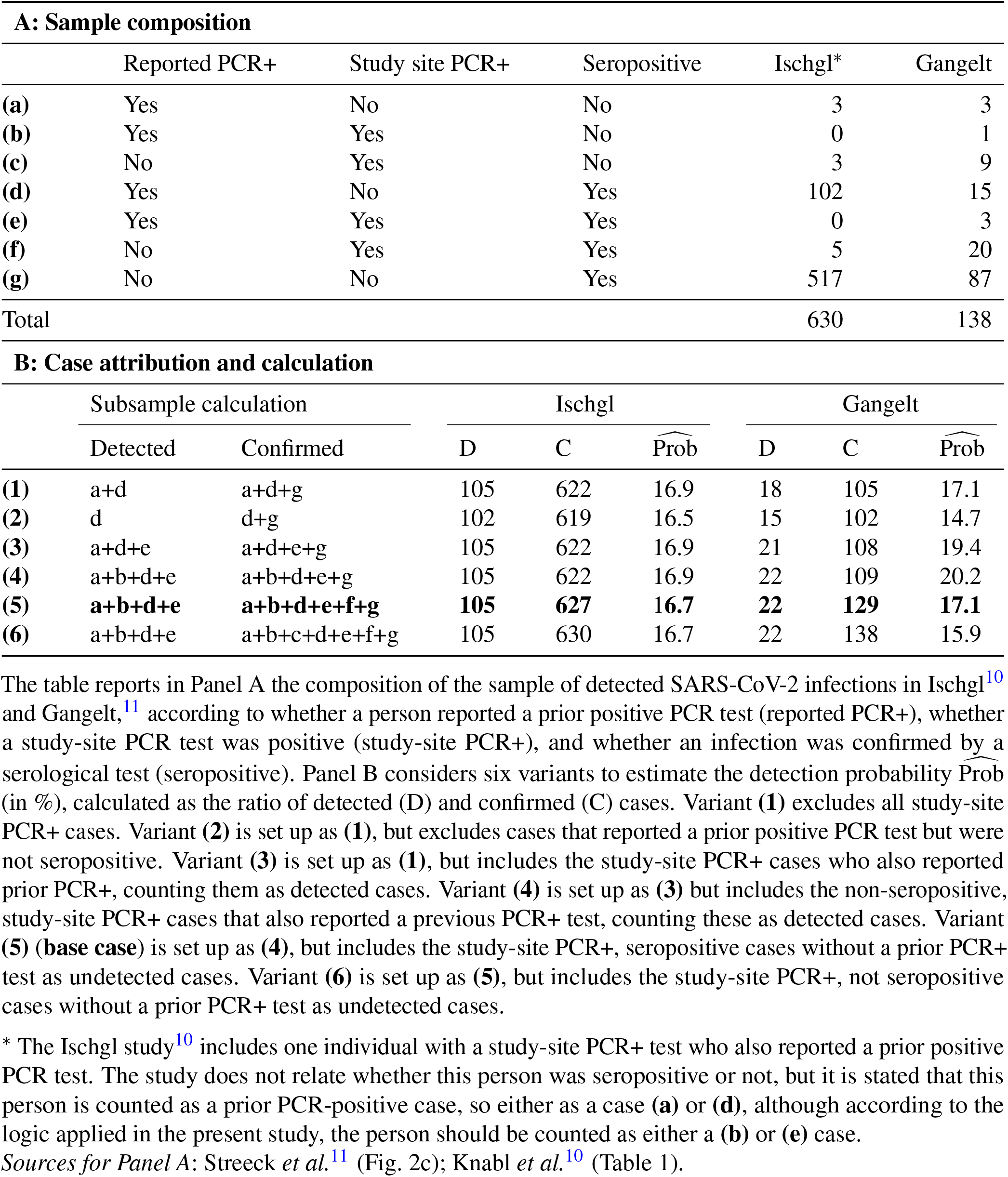
Estimation of the detection probability.

| A: Sample composition |  |  |  |  |  |  |  |  |
| --- | --- | --- | --- | --- | --- | --- | --- | --- |
|  | Reported PCR+ | Study site PCR+ | Seropositive | Ischgl* | Gangelt |  |  |  |
| (a) | Yes | No | No | 3 | 3 |  |  |  |
| (b) | Yes | Yes | No | 0 | 1 |  |  |  |
| (c) | No | Yes | No | 3 | 9 |  |  |  |
| (d) | Yes | No | Yes | 102 | 15 |  |  |  |
| (e) | Yes | Yes | Yes | 0 | 3 |  |  |  |
| (f) | No | Yes | Yes | 5 | 20 |  |  |  |
| (g) | No | No | Yes | 517 | 87 |  |  |  |
| Total |  |  |  | 630 | 138 |  |  |  |
| B: Case attribution and calculation |  |  |  |  |  |  |  |  |
|  | Subsample calculation |  | Ischgl |  |  | Gangelt |  |  |
|  | Detected | Confirmed | D | C | Prob | D | C | Prob |
| (1) | a+d | a+d+g | 105 | 622 | 16.9 | 18 | 105 | 17.1 |
| (2) | d | d+g | 102 | 619 | 16.5 | 15 | 102 | 14.7 |
| (3) | a+d+e | a+d+e+g | 105 | 622 | 16.9 | 21 | 108 | 19.4 |
| (4) | a+b+d+e | a+b+d+e+g | 105 | 622 | 16.9 | 22 | 109 | 20.2 |
| (5) | **a+b+d+e** | **a+b+d+e+f+g** | **105** | **627** | **16.7** | **22** | **129** | **17.1** |
| (6) | a+b+d+e | a+b+c+d+e+f+g | 105 | 630 | 16.7 | 22 | 138 | 15.9 |

The table reports in Panel A the composition of the sample of detected SARS-CoV-2 infections in Ischgl<sup>10</sup> and Gangelt,<sup>11</sup> according to whether a person reported a prior positive PCR test (reported PCR+), whether a study-site PCR test was positive (study-site PCR+), and whether an infection was confirmed by a serological test (seropositive). Panel B considers six variants to estimate the detection probability $\widehat{Prob}$ (in %), calculated as the ratio of detected (D) and confirmed (C) cases. Variant (1) excludes all study-site PCR+ cases. Variant (2) is set up as (1), but excludes cases that reported a prior positive PCR test but were not seropositive. Variant (3) is set up as (1), but includes the study-site PCR+ cases who also reported prior PCR+, counting them as detected cases. Variant (4) is set up as (3) but includes the non-seropositive, study-site PCR+ cases that also reported a previous PCR+ test, counting these as detected cases. Variant (5) (**base case**) is set up as (4), but includes the study-site PCR+, seropositive cases without a prior PCR+ test as undetected cases. Variant (6) is set up as (5), but includes the study-site PCR+, not seropositive cases without a prior PCR+ test as undetected cases.
\* The Ischgl study<sup>10</sup> includes one individual with a study-site PCR+ test who also reported a prior positive PCR test. The study does not relate whether this person was seropositive or not, but it is stated that this person is counted as a prior PCR-positive case, so either as a case (a) or (d), although according to the logic applied in the present study, the person should be counted as either a (b) or (e) case.
Sources for Panel A: Streeck *et al.*<sup>11</sup> (Fig. 2c); Knabl *et al.*<sup>10</sup> (Table 1).

As shown in Table 1, the Ischgl data account for 627 seropositive individuals in the base case specification. Out of these, 105 persons have reported a prior positive PCR test result. Estimating the detection probability by dividing the number of detected cases (prior positive PCR test) by the number of seropositive individuals, the base specification thus yields an estimate of the detection probability of 16.7%. Table 1 shows that, as far as the estimation of the detection probability is concerned, the treatment of the uncertain cases does not matter much. In the six alternative specifications, the detection probability estimates vary from 16.5% to 16.9%.

Table 1 shows that if the information from the Gangelt study is used instead, the effect of the treatment of the uncertain cases has a much greater impact on the estimates of the detection probability. In the Gangelt study, 33 infections were detected by the study-site PCR test, 23 of them were seropositive, ten were not. Of the new PCR-positive cases, four individuals (three seropositive, one not) reported a prior positive PCR test. Table 1 shows that in absolute and relative terms, the number of uncertain cases is notably higher than in the Ischgl study. The Gangelt study reports 102 seropositive individuals who were not study-site PCR-positive. 15 of these reported a prior positive PCR test result. Table 1 shows that the alternative strategies to treat the uncertain cases lead to a wider range of estimates of the detection probability. However, applying the described base case strategy, the estimated detection probability of 17.1% is similar to the Ischgl-based estimate of 16.7%.

Using RKI data with May 1, 2020 as the interval end date combined with the Ischgl data, we obtain a base case IFR estimate of 0.83% with 95% CI [0.69%; 0.98%]. The corresponding estimate using the Gangelt data is 0.85% with 95% CI [0.52%; 1.17%]. The wider confidence bounds are a result of the smaller sample size in the Gangelt study. The Ischgl- and Gangelt-based IFR estimates are not significantly different (p-value: 0.932).

That the IFR estimates based on the results from two independent seroepidemiological studies are similar serves as a first robustness check. Two more are applied. The first consists of a variation of the end date of the time interval used to estimate the CFR based on the RKI data. Figure Fig. 2a shows that the IFR estimate is not sensitive to varying the interval end date from April 1 until June 1, 2020. The IFR estimates also do not change much when modifying the backward-shift of fatalities, which is shown in Figure 3. It depicts the change of the IFR point estimate using the base case end date of May 1, 2020, and where the number of days by which the COVID-19 fatalities are shifted varies from 11 to 27 days. Across the robustness checks (use of Ischgl vs Gangelt study results, variation of the interval end date used to estimate the CFR, and days applied to backward-shift fatalities), the point estimate of the IFR varies in a range from 0.70% to 0.86%.

**Fig. 2.**
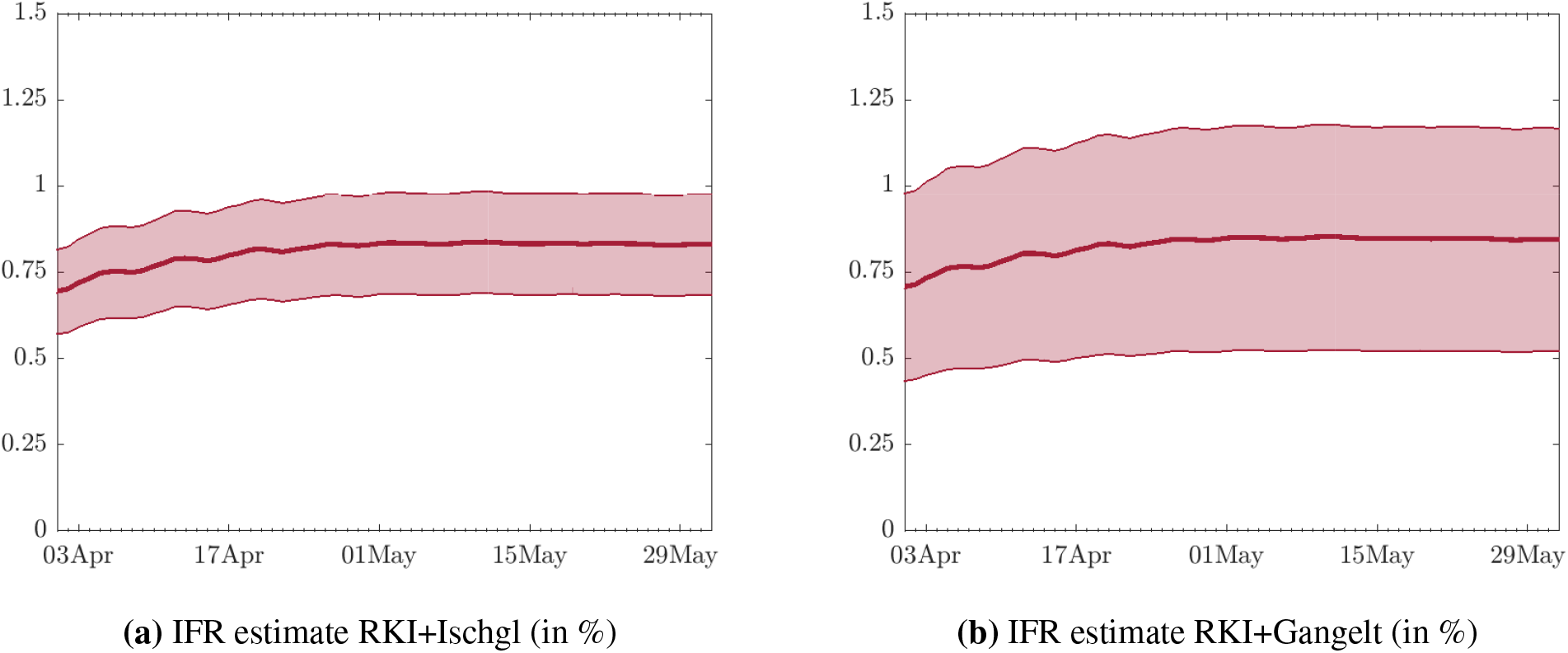
Sensitivity of the IFR estimate with respect to the estimation date of the CFR. The figure presents the two IFR estimates (in %) using data from the RKI combined with Ischgl data (in Panel a) and Gangelt data (in Panel b). The IFR estimate is reported for the period April 1 to June 1, 2020. The shaded areas depict the 95% confidence intervals.

**Fig. 3.**
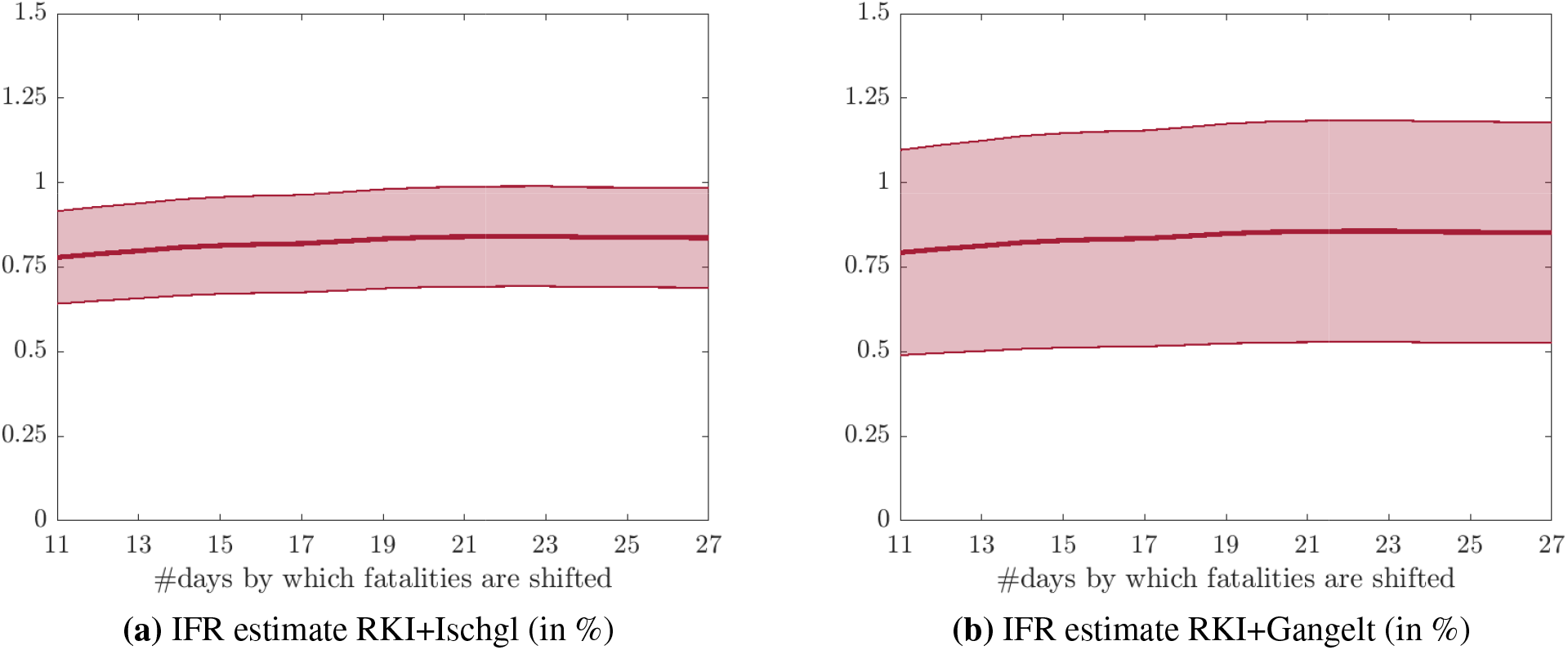
Sensitivity of the IFR estimates with respect to backward shift of COVID-19-associated fatalities. The figure illustrates the sensitivity of the estimated IFR on May 1, 2020, with respect to the number of days by which the number of COVID-19-associated fatalities are shifted backwards. The shaded area depicts the 95% confidence intervals.

## Discussion

In Germany, comprehensive data on detected SARS-CoV-2 infections and COVID-19-associated fatalities are publicly reported by the RKI. Reported infections are detected by SARS-CoV-2 PCR tests, which are, because of limited capacities, predominantly applied to symptomatic patients and their contacts, and members of selected groups, e.g., medical staff. As a result of the selective testing strategy, many asymptomatic SARS-CoV-2 infections or mild courses of COVID-19 are not captured by the data. Since the onset of the pandemic, news about RKI reported SARS-CoV-2 infections and COVID-19 fatalities have consistently been the first topic in the daily news. However, due to the selective testing strategy, the case fatality rate computed based on these data is an unsuitable estimate of the infection fatality rate of SARS-CoV-2 in Germany. Its size, a key indicator of the threat on public health caused by the pandemic, is at the time of writing still a controversially discussed topic, in Germany and elsewhere.

The statistical approach proposed herein relies on the law of total probability to formally relate the CFR and IFR of SARS-CoV-2. This linking formula provides a good approximation of the IFR if a set of assumptions regarding data quality and compatibility as well as sensitivity, specificity, and positive predictive power of the tests employed to detect a SARS-CoV-2 infection hold. The key assumption regarding data quality, which arguably can be maintained for Germany, is that all COVID-19 deaths are reported. It may also be noted that the described selective testing strategy has a favorable impact on the sensitivity and positive predictive value of the selective SARS-CoV-2 PCR test. The linking formula is based on the assumption that these are approximately 100%. A key ingredient of this formula is the detection probability of a SARS-CoV-2 infection. The proposed approach exploits that this probability can be estimated using data obtained by seroepidemiological studies, which use study-site serological tests for antibodies against SARS-CoV-2 combined with PCR tests, and which also collect information about prior positive SARS-CoV-2 PCR tests of the study participants. The seroepidemiological studies conducted in the infection hotspots Ischgl (Austria) and Gangelt (Germany) provide such data. The high specificity of the serological test for antibodies reported in the Ischgl study makes it particularly useful. When combining the country-wide data on SARS-CoV-2 infections and COVID-19 fatalities with data from seroepidemiological studies of infection hotspots, it is also necessary that the testing strategies, country-wide and in the hotspot (prior to the study), correspond, because the linking formula requires that the composition of the populations of infected persons match. Moreover, the composition of the populations at risk of a SARS-CoV-2 infection must be comparable, country-wide and in the study population. The present study uses data from a time period during the first wave of the SARS-CoV-2 pandemic, for which these compatibility assumptions can arguably be defended.

It is obvious that the proposed method must be applied with diligence and accounting for the dynamics of a pandemic. Yet it has certain advantages over extant approaches. In a seroepidemiological study of an infection hotspot, the number of COVID-19-associated fatalities will be small. For example, only two COVID-19-associated deaths were reported in the Ischgl study.^10^ In the municipality of Gangelt, seven COVID-19-associated fatalities where recorded at the time of the study.^11^ The projected number of infected persons in Gangelt was 1,956, resulting in the reported IFR estimate of 0.36% (95% CI: [0.29%; 0.45%]). Following up two weeks beyond the period of the Gangelt study, there is one additional COVID-19-associated death in Gangelt. With eight fatalities, the IFR point estimate increases to 0.41%, and the 95% confidence interval when accounting for uncertainty in the number of deaths is [0.21%; 0.84%].^11^ In fact, the municipality of Gangelt reported nine fatalities associated with COVID-19 on April 24, 2020.^20^ Using the 19 days backward shift of fatalities, the IFR estimate thus increases to 0.46%. Small changes and errors when counting the number of COVID-19-associated fatalities in the infection hotspot can, thus, have a severe impact on the IFR estimate. In the present approach, the number of COVID-19-associated fatalities upon which the IFR estimation is based amounts to 8,059, which helps making the IFR estimate robust towards alternative specifications, and which also enables the use of large sample statistical inference.

The authors of the Gangelt study emphasize that their original IFR estimate should not readily be used to assess the IFR of SARS-CoV-2 in Germany. Notwithstanding, the IFR estimate of 0.36% is frequently used as a reference to gauge the threat that COVID-19 poses to the Germany’s public health, and its relatively small size is put forth as an argument against more austere measures to contain the pandemic. Aside from severe risk group-specific and uncertain long-term effects of COVID-19, which call for a more cautious risk assessment, the present study shows that the IFR of SARS-CoV-2 in Germany may have been notably higher. The base specification IFR estimate of 0.83% (95% CI: [0.69%; 0.98%]), as well as the similar estimates obtained from the robustness checks, correspond more closely to the consolidated estimate of 0.68% (95% CI: [0.53%; 0.82%]) reported in a meta study,^21^ where the difference may reflect the disadvantageous age structure of the German population at risk.

New testing strategies alter the composition of the infected subpopulation, protective measures change the population at risk of a SARS-CoV-2 infection, and progress in medical treatments may hopefully reduce the IFR of SARS-CoV-2. Such dynamics during a pandemic will necessitate a re-assessment of the IFR estimation. When the approach proposed herein is to be used for that purpose, it must be checked whether the assumptions which are necessary for the linking formula to hold can be maintained, and whether and how its elements have to be re-estimated. The data on reported cases and fatalities used to estimate the CFR have to correspond to the pandemic situation, and the results from extant seroepidemiological studies will no longer be useful to estimate the detection probability. When designing a new seroepidemiological study that can be used instead, the following requirements must be met. First, the study should be based on either a random sample or a complete subpopulation. Unfortunately, recent RKI hotspot studies exclude children,^22^ so they are not particularly useful. Second, the testing strategy as it applies to the subpopulation of the serological study, as well as the composition of the study population at risk must match their country-wide counterparts. Third, the seroepidemiological study must inquire about a prior positive PCR test. If these conditions are met, a re-assessment of the IFR becomes possible. These are also the conditions to apply the proposed method to estimate the IFR of SARS-CoV-2 in other (country) populations.

## Methods

### Statistical framework

The framework for statistical modeling employs the following set of assumptions and notation. First, consider a base population of persons (e.g. of a country) who are alive at the onset of the pandemic and at risk of a SARS-CoV-2 infection. A fraction of this population will be infected during a time interval t after the onset of the pandemic. It is assumed that neither the composition of the population at risk nor the strategy to test for SARS-CoV-2 changes during this time interval. The goal is to estimate the infection fatality rate (IFR), which as defined here is the probability that a person in the infected subpopulation (C = 1, else C = 0) dies of or with COVID-19 (M = 1, else M = 0). Such a fatality may occur after the time interval t, but is associated with the infection that happened during t. Moreover, a fraction of the base population is tested for SARS-CoV-2 by a PCR test (T = 1, else T = 0). Implementing the population-wide testing strategy as it applies during the time interval t, this test will not be randomly administered to members of the base population, but applied selectively. Because of limited capacities, testing will focus on symptomatic persons, their contacts, or particular groups. A positive SARS-CoV-2 PCR test is indicated by R = 1, and R = 0 else.

Second, consider a population of persons who take part in a seroepidemiological study in an infection hotspot, and in which a study-site serological test for antibodies against SARS-CoV-2 is performed. The study population can be a random sample,^11^ or the study may target the entire hotspot population.^10^ The composition of the population at risk of a SARS-CoV-2 infection and the selective PCR testing strategy are assumed to correspond in the study and in the base population. Study-site serological testing (and, if applied, study-site PCR testing) is not selective, but administered to the entire study population. For persons who test seropositive (i.e., antibodies against SARS-CoV-2 are detected), R^s^ = 1 (else R^s^ = 0). Moreover, for every member of the study population, information is obtained whether they received a positive SARS-CoV-2 PCR test result before the study (R = 1) or not (R = 0). Seropositive individuals without a prior positive PCR test constitute the group of undetected infections. Finally, the draws from the base population and the study population are assumed to be independent.

In this framework, the probabilities of interest and the assumptions about them are as follows.

a. P(M = 1|C = 1): the infection fatality rate (IFR), estimated.
b. P(R = 1|C = 1): the detection probability, estimated. Note that P(R = 1, T = 1|C = 1) = P(R = 1|C = 1), because R = 1 ⇒ T = 1.
c. P(M = 1|T = 1, R = 1, C = 1): the case fatality rate (CFR), estimated.
d. P(C = 1|T = 1, R = 1): the positive predictive value (PPV) of the selective SARS-CoV-2 PCR test, assumed to be ≈ 1.
e. P(R = 1|T = 1, C = 1): the sensitivity of the selective SARS-CoV-2 PCR test, assumed to be ≈ 1.
f. P(R^s^ = 0| C = 0): the specificity of the study-site serological test, assumed to be ≈ 1. Note that study-site testing includes all study participants.
g. P(M = 1 |T = 0, C = 1): the probability that a COVID-19-associated fatality goes unreported, assumed to be zero.
h. P(T = 1| C = 1): probability that an infected person is tested by the selective PCR test, assumed to be non-zero.
i. P(R^s^ = 1|C = 1): sensitivity of the study-site serological test, assumed to be non-zero.
j. P(C = 1): infection probability, assumed to be non-zero.

By the law of total probability and observing that T = 0 ⇒ R = 0, the IFR can be written as:

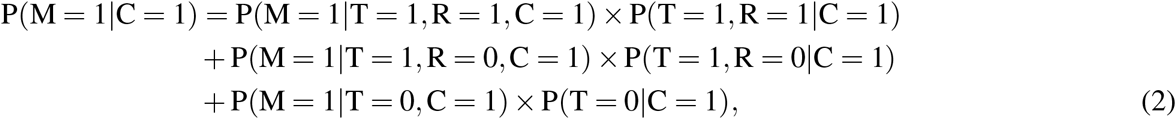

Assumptions (d)-(g) imply that certain terms on the right-hand side of Eq. (2) can be omitted or re-formulated.

i. Assumption (e) implies that P(R = 0 | T = 1, C = 1) ≈ 0, such that P(R = 0, T = 1, C = 1) ≈ 0, provided that P(T = 1, C = 1) *>* 0, which is implied by Assumption (h) and (j). It follows that P(T = 1, R = 0 | C = 1) ≈ 0, such that the second line on the right-hand-side of Eq. (2) is approximately zero and can be omitted.
ii. Assumption (g) together with Assumptions (h) and (j) implies that the third line on the right-hand side of Eq. 2 is zero and can be be omitted.
iii. The detection probability P(T = 1, R = 1 | C = 1) = P(R = 1 | C = 1) on first line of the right hand side of Eq. (2) can be approximated by

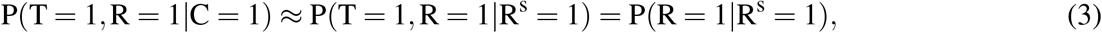

if P(C = 1 | R^s^ = 1) ≈ 1, that is, if the PPV of the study-site serological test is approximately 100%. The PPV close to 100% results if Assumptions (f)), (i) and (j) hold. This claim can be shown by applying Bayes’ law:^23^

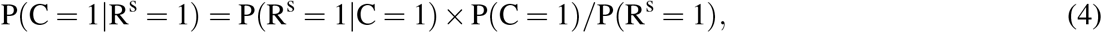

where

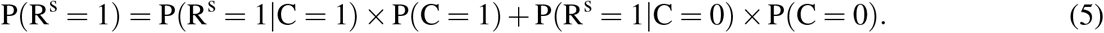 Assumption (f) implies that P(R^s^ = 1|C = 0) ≈ 0. Accordingly, P(C = 1|R^s^ = 1) ≈ 1 if the specificity of the serological test is approximately 100% and P(C = 1) *>* 0 and P(R^s^ = 1|C = 1) *>* 0.
iv. Assumption (e) implies that in the first line of Eq. (2), P(M = 1|R = 1, C = 1) ≈ P(M = 1|R = 1).

Accounting for the implications (i)-(iii), an approximation of the IFR in Eq. (2) can be given as

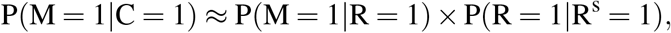

which is the linking formula (1) given in the main text.

### Estimation strategy and large sample statistical inference

Next, consider a strategy to estimate the IFR and to obtain and apply asymptotic inference. For notational brevity, denote P(M = 1 | R = 1) (the CFR) by P_1_, and its estimate by 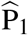. Moreover, denote P(R = 1|R ^s^ = 1) (the detection probability) by P_2_, and its estimate by 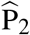. To motivate the estimation strategy and formalize statistical inference, we draw on an approach proposed by Lynch and Wachter^24^ who consider moment-based estimators with varying sample lengths. For that purpose, note that the conditional probabilities P_1_, and P_2_ are also the conditional expected values of the binary random variables M (indicating a COVID-19-associated fatality), and R (indicating a positive selective SARS-CoV-2 PCR test). Using conditional sample means to estimate the conditional expected values thus constitutes a moment-based estimation strategy. It is based on the assumption that the individuals for which we record fatalities are drawn from a population of positively tested individuals whose composition does not change over the time interval t. This is why it is assumed supra that the testing strategy and the composition of the population at risk of a SARS-CoV-2 infection do not change during t. A moment-based estimate of the CFR 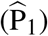 is then obtained by dividing the number of properly back-shifted COVID-19-associated fatalities by N, the number of PCR-positive cases reported in the time interval t. Put differently, the estimate 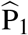 is the sample mean of observations of the random variable M computed on a sample of individuals for which R = 1. Assuming that the conditions to apply a law of large numbers hold, 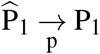 as N → ∞.

A moment-based estimate of the detection probability 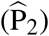 can be calculated by exploiting the information provided by the seroepidemiological study. It is obtained by dividing the number of persons who are both seropositive and (prior to the study) tested PCR-positive by the number of seropositive cases. Put differently, the estimate 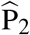 is the mean of observations of the random variable Y computed on a sample of persons for which R^s^ = 1. Let the number of seropositve cases be *λ* N, where 0 *< λ <* 1. Provided that a law of large number holds and assuming that *λ* remains fixed 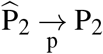 as N → ∞.

An estimate of the IFR is thus obtained by multiplying the CFR estimate with an estimate of the detection probability,

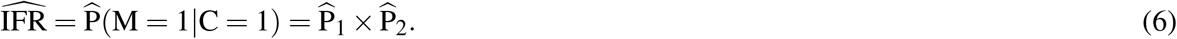

By the continuous mapping theorem 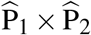 provides a consistent estimate of P_1_×P_2_, the IFR proxy.

In the present case, two seroepidemiological studies (for the infection hotspots Ischgl and Gangelt) can be used for the purpose of estimating the detection probability, which facilitates a specification test. Let the corresponding estimates of the detection probability be denoted 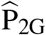, and 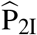 and let *λ*_G_N denote the number of observations used to compute 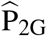, and *λ*_I_N denote the number of observations used to compute 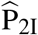. Accordingly, two estimators for the IFR can be considered,

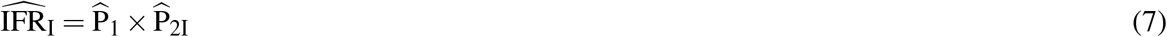

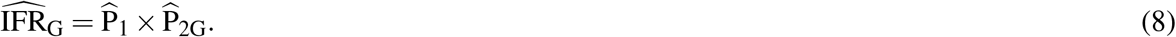

Provided that a law of large numbers holds and *λ*_G_ and *λ*_I_ remain fixed, 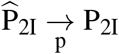and 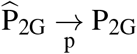 as N→ ∞. If both 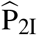 and 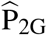 are consistent estimates of the detection probability, then P_2G_ = P_2I_ = P_2_ and the two IFR estimates 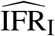 and 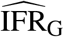 should both converge in probability to the same limit (the IFR proxy).

Large sample inference can be conducted by applying the delta method and accounting for the fact that the samples used to provide the estimates have different lengths^24^. For that purpose, denote 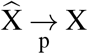 where 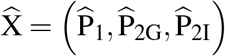 and X = (P_1_, P_2G_, P_2I_)′. Assuming that a central limit theorem applies,

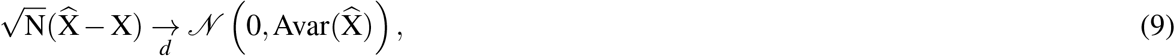

where 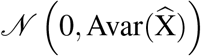 denotes the multivariate normal distribution with mean zero and variance-covariance matrix

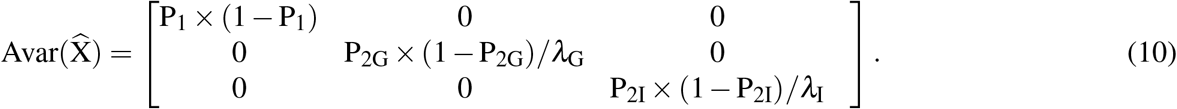

Here, it is assumed that a) as the sample size N grows, the ratio of sample sizes remains the same, that b) the three samples are independently drawn, and c) the observations within the samples are independent, too.

By the continuous mapping theorem, 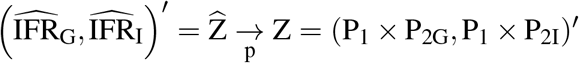. Moreover, applying the delta method gives:

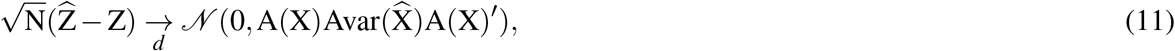

where

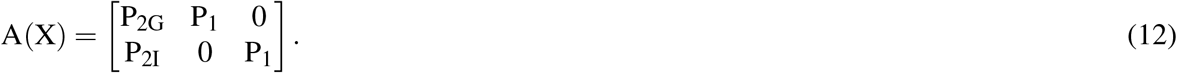

Accordingly, it is possible to use the following approximation of the joint distribution of the two IFR estimates:

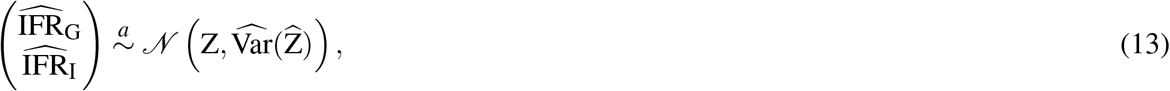

where

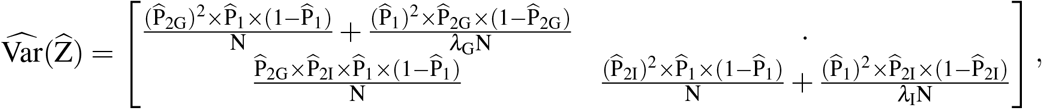

which can be used to compute standard errors and confidence intervals, as well as a Wald-statistic to test the aforementioned hypothesis that P_1_ × P_2G_ = P_1_ × P_2I_.

### Empirical implementation

The empirical analysis is performed in Matlab®, Version 2020b. The Matlab® code is available at https://tinyurl.com/Covid19-IFR-DE.

### Data availability

The data used for the empirical analysis are publicly available from the Robert Koch Institute (https://www.rki.de/DE/Content/InfAZ/N/Neuartiges_Coronavirus/Daten/Fallzahlen_Kum_Tab.html for information on deaths and https://www.rki.de/DE/Content/InfAZ/N/Neuartiges_Coronavirus/Projekte_RKI/Nowcasting.html for the infection nowcasts; both websites were last accessed on December 12, 2020). The data to calculate the probabilities are taken from the seroepidemiological studies in Ischgl^10^ and Gangelt,^11^ respectively. The data are available at https://tinyurl.com/Covid19-IFR-DE.

## Data Availability

The data used for the empirical analysis are publicly available from the Robert Koch Institute
(https://www.rki.de/DE/Content/InfAZ/N/Neuartiges_Coronavirus/Daten/Fallzahlen_Kum_Tab.html for information on deaths and https://www.rki.de/DE/Content/InfAZ/N/Neuartiges_Coronavirus/Projekte_RKI/Nowcasting.html for the infection nowcasts; both websites were last accessed on December 12, 2020). The data to calculate the probabilities are taken from the seroepidemiological studies in
Ischgl (https://www.medrxiv.org/content/10.1101/2020.08.20.20178533v1) and Gangelt (https://doi.org/10.1038/s41467-020-19509-y), respectively.

https://tinyurl.com/Covid19-IFR-DE

## References

1. BMG. Verordnung zum Anspruch auf bestimmte Testungen für den Nachweis des Vorliegens einer Infektion mit dem Coronavirus SARS-CoV-2. Bundesanzeiger AT 09.06.2020 V1, 1–4 (2020).

2. BMG. Verordnung zur Testpflicht von Einreisenden aus Risikogebieten. Bundesanzeiger AT 07.08.2020 V1, 1 (2020).

3. Lai, C.-C. et al.. Asymptomatic carrier state, acute respiratory disease, and pneumonia due to severe acute respiratory syndrome coronavirus 2 (SARS-CoV-2): Facts and myths. J. microbiology, immunology, infection 53, 404–412, DOI: https://doi.org/10.1016/j.jmii.2020.02.012 (2020).

4. Lau, H. et al.. Internationally lost COVID-19 cases. J. Microbiol. Immunol. Infect. 53, 454 – 458, DOI: https://doi.org/10.1016/j.jmii.2020.03.013 (2020).

5. Gao, Z. et al.. A systematic review of asymptomatic infections with COVID-19. J. Microbiol. Immunol. Infect. DOI: https://doi.org/10.1016/j.jmii.2020.05.001 (2020).

6. Götzinger, F. et al.. COVID-19 in children and adolescents in Europe: a multinational, multicentre cohort study. The Lancet Child & Adolesc. Heal. 4, 653 – 661, DOI: https://doi.org/10.1016/S2352-4642(20)30177-2 (2020).

7. Yasuhara, J., Kuno, T., Takagi, H. & Sumitomo, N. Clinical characteristics of COVID-19 in children: A systematic review. Pediatr. Pulmonol. 55, 2565–2575, DOI: https://doi.org/10.1002/ppul.24991 (2020). https://onlinelibrary.wiley.com/doi/pdf/10.1002/ppul.24991.

8. RKI. Täglicher Lagebericht des RKI zur Coronavirus-Krankheit-2019 (COVID-19). Tech. Rep., Robert Koch Institut (2020). Available at https://www.rki.de/DE/Content/InfAZ/N/Neuartiges_Coronavirus/Situationsberichte/2020-08-04-en.pdf.

9. Ouslander, J. G. & Grabowski, D. C. Covid-19 in nursing homes: Calming the perfect storm. J. Am. Geriatr. Soc. 68, 2153–2162, DOI: https://doi.org/10.1111/jgs.16784 (2020). https://agsjournals.onlinelibrary.wiley.com/doi/pdf/10.1111/jgs.16784.

10. Knabl, L. et al.. High SARS-CoV-2 seroprevalence in children and adults in the Austrian ski resort Ischgl (2020). Available at https://www.medrxiv.org/content/10.1101/2020.08.20.20178533v1.

11. Streeck, H. et al.. Infection fatality rate of SARS-CoV2 in a super-spreading event in Germany. Nat. Commun. 11, DOI: https://doi.org/10.1038/s41467-020-19509-y (2020).

12. Robert Koch Institut. Bericht zur Optimierung der Laborkapazitäten zum direkten und indirekten Nachweis von SARS-CoV-2 im Rahmen der Steuerung von Maßnahmen (2020). Available at https://www.rki.de/DE/Content/InfAZ/N/Neuartiges_Coronavirus/Laborkapazitaeten.pdf? blob=publicationFile.

13. Foundation for Innovative New Diagnostics. FIND evaluation update: SARS-CoV-2 molecular diagnostics (2020). Available at https://www.finddx.org/covid-19/sarscov2-eval-molecular/ [Accessed Dec 10, 2020].

14. Backer, J. A., Klinkenberg, D. & Wallinga, J. Incubation period of 2019 novel coronavirus (2019-nCoV) infections among travellers from Wuhan, China, 20–28 January 2020. Euro Surveillance 25, DOI: https://doi.org/10.2807/1560-7917.ES.2020.25.5.2000062 (2020).

15. Böhmer, M. M. et al.. Investigation of a COVID-19 outbreak in Germany resulting from a single travel-associated primary case: a case series. Lancet 20, 920–928, DOI: https://doi.org/10.1016/S1473-3099(20)30314-5 (2020).

16. Verity, R. et al.. Estimates of the severity of coronavirus disease 2019: a model-based analysis. The Lancet Infect. Dis. 20, 669 – 677, DOI: https://doi.org/10.1016/S1473-3099(20)30243-7 (2020).

17. Wang, W., Tang, J. & Wei, F. Updated understanding of the outbreak of 2019 novel coronavirus (2019-nCoV) in Wuhan, China. J. Med. Virol. 92, 441–447, DOI: https://doi.org/10.1002/jmv.25689 (2020).

18. Mimkes, J. & Janssen, R. Test-adjusted results of mortality for Covid-19 in Germany, USA, UK (2020). Available at https://www.medrxiv.org/content/early/2020/11/04/2020.11.03.20225268.

19. Arentz, C. & Wild, F. Vergleich europäischer Gesundheitssysteme in der Covid-19-Pandemie. Tech. Rep. 3/2020, WIP – Wissenschaftliches Institut der PKV, Köln (2020).

20. Gemeinde Gangelt. Coronavirus im Kreis Heinsberg: Stand 24.04.2020 (2020). Available at https://www.gangelt.de/news/226-erster-corona-fall-in-nrw.

21. Meyerowitz-Katz, G. & Merone, L. A systematic review and meta-analysis of published research data on COVID-19 infection-fatality rates. medRxiv DOI: https://doi.org/10.1101/2020.05.03.20089854 (2020).

22. RKI. Studie CORONA-MONITORING lokal (2020). https://www.rki.de/DE/Content/Gesundheitsmonitoring/Studien/cml-studie/cml-studie_node.html [Accessed Jan 25, 2021].

23. Manski, C. F. & Molinari, F. Estimating the COVID-19 infection rate: Anatomy of an inference problem. J. Econom. DOI: https://doi.org/10.1016/j.jeconom.2020.04.041 (2020).

24. Lynch, A. W. & Wachter, J. A. Using samples of unequal length in generalized method of moments estimation. The J. Financial Quant. Analysis 48, 277–307, DOI: https://doi.org/10.1017/S0022109013000070 (2013).

